# Reliability of multi-modal MRI-derived brain phenotypes for multi-site assessment of neuropsychiatric complications of SARS-CoV-2 infection

**DOI:** 10.1101/2021.10.13.21264967

**Authors:** Eugene Duff, Fernando Zelaya, Fidel Alfaro Almagro, Karla L Miller, Naomi Martin, Thomas E. Nichols, Bernd Taschler, Ludovica Griffanti, Christoph Arthofer, Chaoyue Wang, Richard A.I. Bethlehem, Klaus Eickel, Matthias Günther, David K Menon, Guy Williams, Bethany Facer, Greta K Wood, David J Lythgoe, Flavio Dell’Acqua, Steven CR Williams, Gavin Houston, Simon Keller, Gerome Breen, Benedict D Michael, Peter Jezzard, Stephen M Smith, Edward T. Bullmore, On behalf of the COVID-CNS Consortium

**Author notes:** Correspondence: Edward Bullmore, Eugene Duff.

## Abstract

**Background:** Magnetic resonance imaging (MRI) of the brain could be a key diagnostic and research tool for understanding the neuropsychiatric complications of COVID-19. For maximum impact, multi-modal MRI protocols will be needed to measure the effects of SARS-CoV2 infection on the brain by diverse potentially pathogenic mechanisms, and with high reliability across multiple sites and scanner manufacturers.

**Methods:** A multi-modal brain MRI protocol comprising sequences for T1-weighted MRI, T2-FLAIR, diffusion MRI (dMRI), resting-state functional MRI (fMRI), susceptibility-weighted imaging (swMRI) and arterial spin labelling (ASL) was defined in close approximation to prior UK Biobank (UKB) and C-MORE protocols for Siemens 3T systems. We iteratively defined a comparable set of sequences for General Electric (GE) 3T systems. To assess multi-site feasibility and between-site variability of this protocol, N=8 healthy participants were each scanned at 4 UK sites: 3 using Siemens PRISMA scanners (Cambridge, Liverpool, Oxford) and 1 using a GE scanner (King’s College London). Over 2,000 Imaging Derived Phenotypes (IDPs) measuring both data quality and regional image properties of interest were automatically estimated by customised UKB image processing pipelines. Components of variance and intra-class correlations were estimated for each IDP by linear mixed effects models and benchmarked by comparison to repeated measurements of the same IDPs from UKB participants.

**Results:** Intra-class correlations for many IDPs indicated good-to-excellent between-site reliability. First considering only data from the Siemens sites, between-site reliability generally matched the high levels of test-retest reliability of the same IDPs estimated in repeated, within-site, within-subject scans from UK Biobank. Inclusion of the GE site resulted in good-to-excellent reliability for many IDPs, but there were significant between-site differences in mean and scaling, and reduced ICCs, for some classes of IDP, especially T1 contrast and some dMRI-derived measures. We also identified high reliability of quantitative susceptibility mapping (QSM) IDPs derived from swMRI images, multi-network ICA-based IDPs from resting-state fMRI, and olfactory bulb structure IDPs from T1, T2-FLAIR and dMRI data.

**Conclusion:** These results give confidence that large, multi-site MRI datasets can be collected reliably at different sites across the diverse range of MRI modalities and IDPs that could be mechanistically informative in COVID brain research. We discuss limitations of the study and strategies for further harmonization of data collected from sites using scanners supplied by different manufacturers. These protocols have already been adopted for MRI assessments of post-COVID patients in the UK as part of the COVID-CNS consortium.

## Introduction

It is increasingly clear that systemic infection with severe acute respiratory syndrome coronavirus 2 (SARS-CoV-2) is often associated with acute neurological complications at the time of infection, as well as post-acute neurological, cognitive and mental health sequelae that can persist for at least 6 months after infection(Paterson et al., 2020). It seems likely that SARS-CoV-2 infection can have adverse effects on healthy brain function and structure that account for its broad spectrum of neuropsychiatric complications. The causal or pathogenic mechanisms are not yet defined but are likely to be several, including at least (i) viral infection of the central nervous system (CNS), (ii) host immune response to infection, and (iii) cerebrovascular disruption. For precisely targeted interventions, it will be important to know which pathogenic mechanisms are most relevant for which individual patients, or for which syndromically typical groups of patients.

Magnetic resonance imaging (MRI) could be a key diagnostic tool in understanding the impacts of systemic SARS-CoV2 infection on the brain and advancing to better treatments for neuropsychiatric complications of COVID-19 in future. Large-scale post-COVID MRI databases will be important because of the geographic, demographic and clinical heterogeneity of neurological, mental health and cognitive syndromes that have been reported as acute or post-acute outcomes of SARS-CoV-2 infection. To acquire such databases requires multi-modal acquisition protocols and analysis pipelines that can be reliably implemented across a variety of scanner manufacturers and models. Ideally, multi-modal MRI protocols for post-COVID research should also be well matched to existing large-scale neuroimaging databases with relevant demographic profiles, such as the UK Biobank database of adults with mean age of 50 years. Here we describe the technical development and validation by a “travelling heads” study of a multi-site protocol for the COVID-CNS consortium, which aims to collect data on ∼700 post-COVID neurological cases and controls from a national network of UK sites.

We started from the principle that a standard brain MRI protocol, robust enough to be reliably implemented across multiple sites and scanners, should also be inclusive of different modalities of MRI that can provide distinct or complementary insights into candidate pathogenic mechanisms. For example, the C-MORE consortium for multi-organ MRI studies of post-hospitalised COVID cases (3) has used a set of 7 brain MRI sequences (**Table 1**) to measure T1-weighted MRI, T2-FLAIR, diffusion MRI (dMRI), susceptibility-weighted MRI (swMRI), and arterial spin labelling (ASL). The inclusion of each of these sequences was justified by their diagnostic relevance to distinct pathogenic mechanisms: e.g., swMRI is a marker of iron deposition and micro-haemorrhages and ASL measures parameters of regional cerebral blood flow, so both are relevant to vascular mechanisms; T2-FLAIR is a widely used measure of inflammation-related changes in white matter; T1- and dMRI-derived brain structural phenotypes have been found to be associated with immune cell counts in blood samples from post-COVID patients (Griffanti et al., 2021). T1-weighted data have also been used to measure volume and tissue contrast of the olfactory bulb and brain stem structures that are most likely to be neurotropically infected via olfactory nerve terminals and other specialist sensory receptors. Thus, the inclusion of sequences in the C-MORE neuro-MRI protocol was well-motivated; but the requirement to complete all neuroimaging sequences in less than 20 mins, as part of a 70 min multi-organ MRI protocol, meant that some potentially informative sequences were excluded (fMRI) or abbreviated (dMRI, ASL).

**Table 1:**
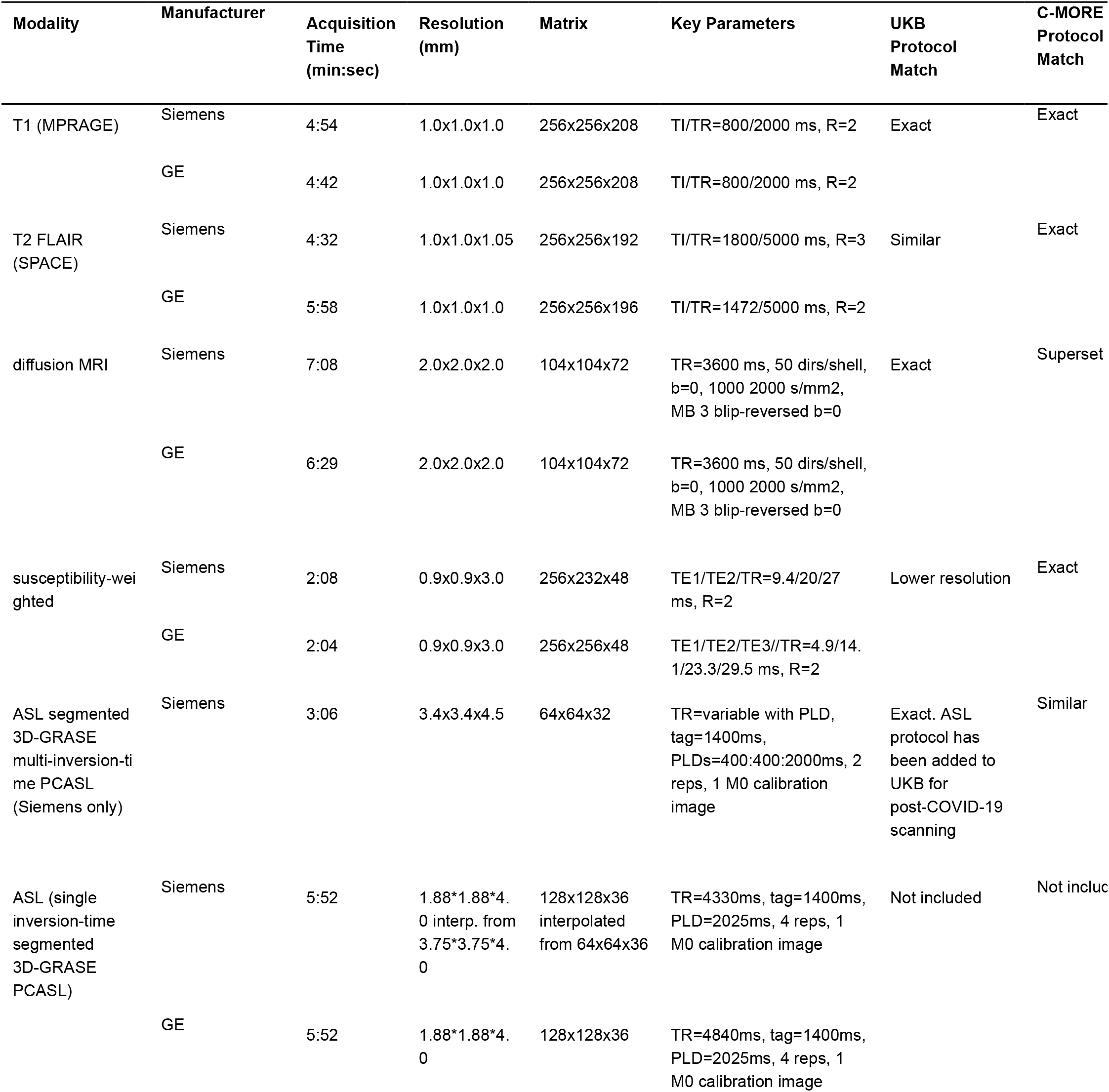
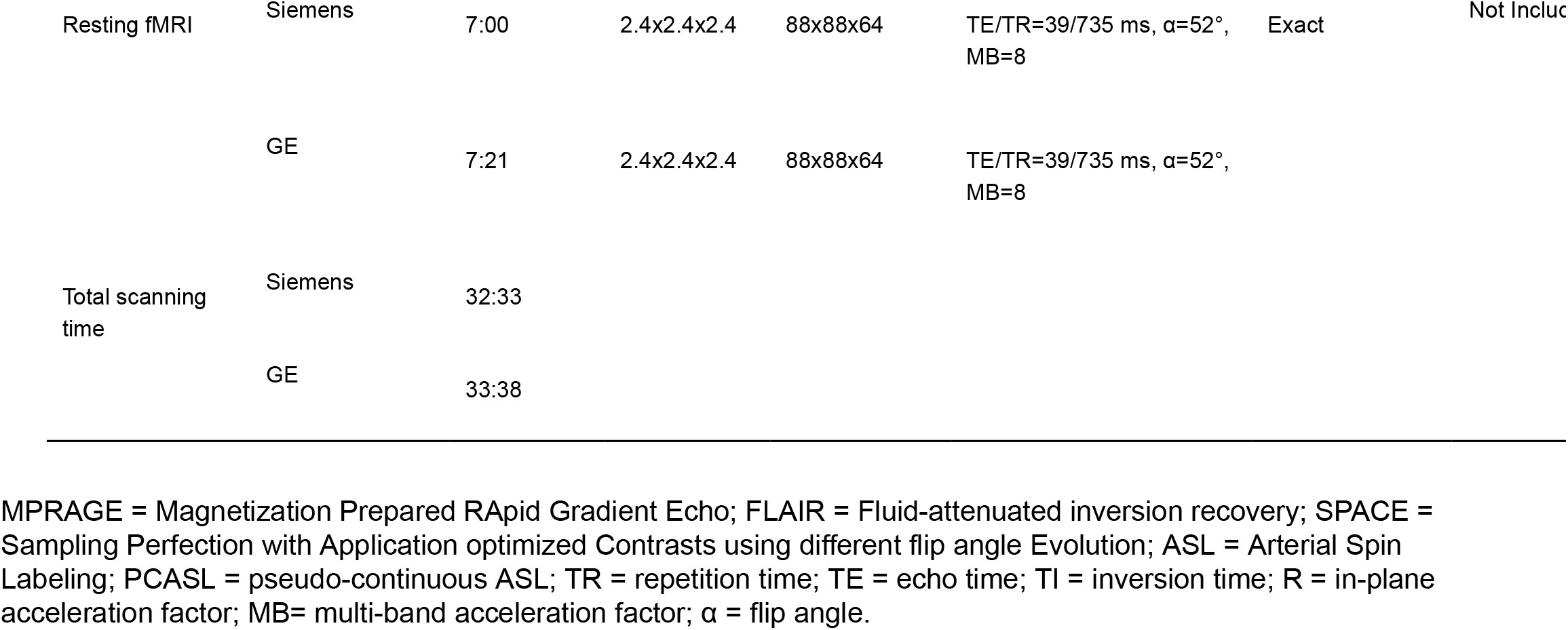
Multimodal MRI protocols for COVID-related neuroimaging with Siemens and GE 3T scanners.

| Modality | Manufacturer | Acquisition Time (min:sec) | Resolution (mm) | Matrix | Key Parameters | UKB Protocol Match | C-MORE Protocol Match |
| --- | --- | --- | --- | --- | --- | --- | --- |
| T1 (MPRAGE) | Siemens | 4:54 | 1.0x1.0x1.0 | 256x256x208 | TI/TR=800/2000 ms, R=2 | Exact | Exact |
|  | GE | 4:42 | 1.0x1.0x1.0 | 256x256x208 | TI/TR=800/2000 ms, R=2 |  |  |
| T2 FLAIR (SPACE) | Siemens | 4:32 | 1.0x1.0x1.05 | 256x256x192 | TI/TR=1800/5000 ms, R=3 | Similar | Exact |
|  | GE | 5:58 | 1.0x1.0x1.0 | 256x256x196 | TI/TR=1472/5000 ms, R=2 |  |  |
| diffusion MRI | Siemens | 7:08 | 2.0x2.0x2.0 | 104x104x72 | TR=3600 ms, 50 dirs/shell, b=0, 1000 2000 s/mm<sup>2</sup>, MB 3 blip-reversed b=0 | Exact | Superset |
|  | GE | 6:29 | 2.0x2.0x2.0 | 104x104x72 | TR=3600 ms, 50 dirs/shell, b=0, 1000 2000 s/mm<sup>2</sup>, MB 3 blip-reversed b=0 |  |  |
| susceptibility-weighted | Siemens | 2:08 | 0.9x0.9x3.0 | 256x232x48 | TE1/TE2/TR=9.4/20/27 ms, R=2 | Lower resolution | Exact |
|  | GE | 2:04 | 0.9x0.9x3.0 | 256x256x48 | TE1/TE2/TE3/TR=4.9/14.1/23.3/29.5 ms, R=2 |  |  |
| ASL segmented 3D-GRASE multi-inversion-time PCASL (Siemens only) | Siemens | 3:06 | 3.4x3.4x4.5 | 64x64x32 | TR=variable with PLD, tag=1400ms, PLDs=400:400:2000ms, 2 reps, 1 M0 calibration image | Exact. ASL protocol has been added to UKB for post-COVID-19 scanning | Similar |
| ASL (single inversion-time segmented 3D-GRASE PCASL) | Siemens | 5:52 | 1.88*1.88*4.0 interp. from 3.75*3.75*4.0 | 128x128x36 interpolated from 64x64x36 | TR=4330ms, tag=1400ms, PLD=2025ms, 4 reps, 1 M0 calibration image | Not included | Not included |
|  | GE | 5:52 | 1.88*1.88*4.0 | 128x128x36 | TR=4840ms, tag=1400ms, PLD=2025ms, 4 reps, 1 M0 calibration image |  |  |
| Resting fMRI | Siemens | 7:00 | 2.4x2.4x2.4 | 88x88x64 | TE/TR=39/735 ms, $\alpha=52^\circ$, MB=8 | Exact | Not Included |
| | GE | 7:21 | 2.4x2.4x2.4 | 88x88x64 | TE/TR=39/735 ms, $\alpha=52^\circ$, MB=8 | | |
| Total scanning time | Siemens | 32:33 |  |  |  |  |  |
|  | GE | 33:38 |  |  |  |  |  |

MPRAGE = Magnetization Prepared RAPid Gradient Echo; FLAIR = Fluid-attenuated inversion recovery; SPACE = Sampling Perfection with Application optimized Contrasts using different flip angle Evolution; ASL = Arterial Spin Labeling; PCASL = pseudo-continuous ASL; TR = repetition time; TE = echo time; TI = inversion time; R = in-plane acceleration factor; MB= multi-band acceleration factor; $\alpha$ = flip angle.

In this context, we designed a multi-modal MRI protocol specifically for neuroimaging of post-COVID cases. To optimise comparability with data collected by UKB and C-MORE protocols, we selected Siemens 3T sequences that were as close as possible to these standards, including a multiband sequence for resting state fMRI (implemented in UKB but not in C-MORE) and increasing the scanning time for dMRI and ASL sequences to improve data quality compared to C-MORE. We also iteratively defined a set of General Electric (GE) 3T sequences that approximated as closely as possible the parameters of the Siemens sequences (**Table 1**). Based on our clinical experience to date (2), we rationed the total scanning time of all sequences combined to 30 mins, expecting this to require less than 40 mins of in-scanner time for patients to complete.

To assess the multi-site feasibility and between-site reliability of these protocols, we conducted a “travelling heads” experiment(Weiskopf et al., 2013) whereby N=8 healthy volunteers were scanned once at each of 4 UK sites: 3 using Siemens PRISMA 3T systems (Cambridge, Liverpool and Oxford) and 1 using a GE MR750 Discovery 3T system (King’s College London). Multi-site consistency of neuroimaging data was evaluated along several domains including quality control (QC) criteria, tissue contrast metrics, and multiple categories of imaging-derived phenotypes (IDPs), estimated using customised UKB image-processing pipelines. Linear mixed effects models were used to estimate components of variance and intra-class correlation coefficients as measures of between-site reliability for each metric and IDP. We focus specifically on two questions of interest: (i) How does between-site and between-manufacturer reliability of multi-modal IDPs estimated from these data compare to the benchmark of test-retest reliability of IDPs estimated from repeated scans of UKB participants using a Siemens SKYRA system? (ii) Which are the most (and least) reliable of the thousands of IDPs that can be measured in these data?

## Methods

### Study design and sample

The “travelling heads” design followed previous studies for evaluation of multi-site MRI protocols (3). Each of N=8 healthy participants (7F, age range 21-37 y) was scanned 4 times, once at each of the 4 pilot sites: the Wolfson Brain Imaging Centre at the University of Cambridge; the Wellcome Trust-National Institute of Health Research Clinical Research Facility at King’s College Hospital, King’s College London (KCL); the Liverpool Magnetic Resonance Imaging Centre (LiMRIC) at the University of Liverpool; and the Wellcome Centre for Integrative Neuroimaging at the University of Oxford.

Due to lockdown restrictions prevailing in the UK at the time of scanning (Dec 2020 – Feb 2021), all participants were recruited at one site (KCL) and the ordering and timing of safe travel to other sites was decided pragmatically. Participants were paid an honorarium to compensate for the time taken to complete the protocol. All participants gave informed consent in writing and the study was approved by the Human Biology Research Ethics Committee, University of Cambridge (HBREC.2020.44).

### Scanners and scanning sequences

The Cambridge, Liverpool and Oxford sites all used 3T MAGNETOM PRISMA MRI systems (Siemens Healthineers, Erlangen, Germany) fitted with a 32 channel, receive-only head coil. KCL used a 3T General Electric MR 750 Discovery MRI scanner (GE Healthcare, Waukesha, Wisconsin, USA) and a 32-channel, receive-only head coil (Nova Medical, Wilmington, Massachusetts, USA).

The 3 Siemens scanners implemented the set of 8 sequences summarised in **Table 1**. The sequence for T1-weighting was implemented identically across UKB, C-MORE and COVID-CNS protocols. dMRI and fMRI were implemented in COVID-CNS exactly as in the UKB protocol (the C-MORE protocol included a shorter dMRI sequence and did not include fMRI). T2 FLAIR and swMRI sequences were slightly modified from UKB standards in order to more closely match corresponding sequences in the C-MORE protocol. A multi-post label delay (PLD) 3D-GRASE ASL sequence(Günther et al., 2005) was used identically to that planned to be adopted by UKB COVID study (Douaud et al., 2021) (different to the 2D multi-slice sequence used in C-MORE); a single delay ASL sequence was additionally used to match the ASL imaging pulse sequence of the GE scanner.

The GE scanner implemented an analogous set of 8 sequences (**Table 1**). In most cases it was possible to approximate the parameters of the Siemens sequences by bespoke programming of the default GE sequences for T1-weighted, T2-FLAIR, dMRI, swMRI and fMRI. The GE scanner could not implement the Siemens multi-post label delay ASL sequence with sufficient similarity to the Siemens implementation; so a single post label delay sequence was used for ASL on the GE platform.

### Image processing pipelines and IDPs

Each MRI modality was analysed using custom pipelines for image pre-processing and estimation of multiple MRI contrast metrics and imaging-derived phenotypes (IDPs) derived from the UKB analysis pipelines (www.fmrib.ox.ac.uk/ukbiobank/) (Alfaro-Almagro et al., 2018) and software tools from the FMRIB Software Library (Jenkinson et al., 2012). Pipeline changes were implemented to accommodate minor differences in imaging parameters between UKB and COVID-CNS protocols, to analyse MRI modalities not included in the UKB MRI protocol, e.g., ASL, and to analyse MRI data acquired using the GE scanner at KCL. Where protocols matched exactly, analysis pipelines were identical to those used in the C-MORE COVID study (Griffanti et al., 2021; Raman et al., 2021). Summaries of pre-processing and IDP estimation are provided below for individual modalities, with further details available in (Douaud et al., 2021; Griffanti et al., 2021). For presentation, IDPs reflecting the same phenotypic properties were grouped together into IDP classes(Douaud et al., 2021; Elliott et al., 2018).

### T1-weighted and T2-FLAIR

Processing of T1-weighted and T2-FLAIR data included removal of the face, brain extraction, and registration to the MNI152 brain template (Jenkinson 2002, Andersson 2008). We measured spatial signal-to-noise ratio (SNR) and grey/white contrast-to-noise ratio (CNR) as quality control (QC) metrics. As the T1-weighted image was the primary modality for inter-subject registrations, we also measured QC metrics of registration quality. For Siemens scanners we used an in-house 3d gradient distortion correction developed for the UK Biobank and Human Connectome Project (Alfaro-Almagro et al., 2018), while for the GE site, standard GE gradient distortion correction was implemented. FAST was used to segment images into grey matter, white matter, and cerebro-spinal fluid (Zhang 2001). SIENAX (Smith, 2002) was used to estimate volume measures from these segmentations. Grey matter volumes were estimated for each of 139 regions of interest (ROIs) defined by the Harvard-Oxford cortical and subcortical atlases and the Diedrichsen cerebellar atlas. Sub-cortical volumes were estimated utilizing population priors on shape and intensity variation across subjects (Patenaude et al., 2011). Using an additional non-linear registration procedure, regional volumes of the olfactory bulbs were estimated using T1-weighted, T2-FLAIR and dMRI data, and a template derived from over 700 UKB individuals (Arthofer et al., 2021; Griffanti et al., 2021; Lange et al., 2020).

T2-FLAIR pre-processing was very similar to the T1w pipeline (with the T1-weighted image used for registration to the MNI standard template). Images were segmented using BIANCA to identify white matter (WM) hyperintensities (WMH) (Griffanti et al., 2016), using the UKB BIANCA training file. Periventricular WMH (pWMH) and deep WMH (dWMH) volumes were defined for complementary subsets of total WM hyperintensities that were, respectively, less than (or more than) 10 mm distant from the lateral ventricles (Griffanti et al., 2021).

T1-weighted and T2-FLAIR images were combined in FreeSurfer to model the cortical surface (Desikan et al., 2006; Fischl et al., 2004). This analysis produced IDPs encompassing metrics of subcortical segmentation, regional surface area, volume and mean cortical thickness from a number of different parcellations, and grey-white intensity contrasts (expressed as the fractional contrast between white and grey matter intensities as sampled either side of the grey-white cortical boundary)(Smith et al., 2020). In total 1073 IDPs were measured from T1w and T2_FLAIR scans.

#### swMRI

For the Siemens sequence, the magnitude images from the two echoes of the swMRI data were combined to provide a mapping of T_2_^*^ signal decay. Median T2^*^ was calculated for 14 subcortical structures defined by registration with the parcellated T1 data. To enable qualitative neurological assessment of individual patients the median phase and magnitude data were processed to provide maps highlighting features indicative of abnormal iron deposition, e.g., due to microbleeds. Quantitative susceptibility mapping (QSM) was also performed, using the phase data and a recently developed UKB pipeline (Wang et al., 2021). Susceptibility maps were generated using the iLSQR algorithm (Li et al., 2015), with susceptibility values reported relative to the susceptibility measured in CSF. For the GE sequence, swMRI data had a different number of echoes and required adjusted procedures. In total 28 IDPs were measured from swMRI scans.

#### ASL

For the Siemens sequence, we used the BASIL tools in FSL to estimate maps of cerebral blood flow (CBF) from single-PLD data and CBF and arterial transit time (ATT) from multi-PLD data. BASIL analysis included motion correction and distortion correction using the blip up/down dMRI data. Label and control images were subtracted and a kinetic model was fitted with modelling of the macrovascular component. The M0 calibration image acquired without ASL preparation was used to quantify CBF. Tissue-specific CBF was achieved by projecting grey and white partial volume maps from the T1w image segmented by FAST into the ASL native space.

Grey and white matter masks were defined using partial volume thresholds of 50% and 80% respectively. To avoid dependence on site-specific T1w data, we used T1w data from all sites to define generic masks for estimation of mean grey matter CBF and ATT. In total 4 IDPs were measured from both the multi- and single-PLD ASL data. fMRI: For the Siemens sequence, the multiband-8 fMRI data were corrected for gradient and EPI distortions, motion-corrected using linear alignment using the UKB Resting fMRI pipeline (Alfaro-Almagro et al., 2018), and aligned to the T1w image via a single-band reference image. For the GE sequence the first high-contrast fMRI image prior to magnetisation stabilisation was used for T1w registration. FIX ICA-based denoising was applied using the UKB training dataset (Salimi-Khorshidi et al., 2014). Two sets of resting-state networks derived from group ICA decompositions of UKB reference data (25 and 100 component decompositions) were projected onto the pre-processed resting state fMRI data in a dual-regression analysis (Nickerson et al., 2017). Two whole brain functional connectivity matrices were compiled from all possible partial correlations, and the amplitudes (standard deviations) of spontaneous activity at each regional node were estimated (Alfaro-Almagro et al., 2018). As individual connections showed low test-retest reliability in the UKB dataset, we used a dimension-reduction approach which applied ICA to all functional connectivity IDPs to produce 6 primary modes of variation (Elliott et al., 2018).

These six modes were projected onto the individual’s functional connectivity matrix and used as additional IDPs. In total 3464 IDPs were measured from the fMRI data.

#### dMRI

For the Siemens and GE sequences, dMRI data were closely matched to the UKB sequence and processed using UKB pipelines with minimal alterations (Alfaro-Almagro et al., 2018). The AP-encoding data were pre-processed to remove effects of eddy currents, head motion, and slice dropouts, followed by gradient distortion correction. DTIFIT used the b=1000 shell for diffusion tensor image fitting (Basser et al., 1994) to estimate parameters including fractional anisotropy (FA), tensor mode (MO) and mean diffusivity (MD). The multi-shell data were processed with NODDI (Neurite Orientation Dispersion and Density Imaging) (Zhang et al., 2012), to produce microstructural parameters including ICVF (intra-cellular volume fraction - an index of white matter neurite density), ISOVF (isotropic or free water volume fraction), and ODI (orientation dispersion index, a measure of within-voxel tract disorganisation). These parameters were summarised using two approaches: first using a white-matter tract skeleton analysis producing average values for 48 standard-space tract masks (Smith et al., 2006); and second using probabilistic tractography to provide weighted-mean summaries of the parameters for 27 major tracts. In total 675 IDPs were measured from the dMRI data.

### Statistical analysis and UK Biobank benchmarking

Site and scanner manufacturer can affect the distribution of phenotypes derived from brain images, adding variability and reducing experimental power in multi-site studies. Site effects limited to location shifts and scale changes are easily modeled if they can be estimated, and will result in subject ranking being preserved across sites. Here we characterise the effects of site on the location and scale of IDPs, and compare intra-class correlations (ICCs) of IDPs measured 4 times for each subject scanned at 4 different sites in the travelling heads study, against ICCs of the same IDPs measured twice for each subject (with a 2y interval) at the same site as part of the longitudinal data previously acquired as part of of the UKB imaging enhancement programme (Littlejohns et al., 2020).

Location effects were assessed using repeated-measures ANOVAs, with sphericity assessed using Mauchly’s test. Site-specific means and sphericity tests were computed for all IDPs. We tested the set of null hypotheses that there is zero between-site difference in mean, and the null hypothesis of sphericity for each of 2258 (total) IDPs (excluding IDPs representing individual functional network connections), setting the threshold for refutation of the null by the false discovery rate (FDR=5%, within each type of test), to control type 1 errors in the context of multiple testing entailed by regional resolution of multi-modal MRI. Site-specific effects on each IDP were estimated twice: once using all the analysable data (from 4 sites, including 1 GE site), and once using only Siemens data (from 3 sites). This allowed us to investigate site-differences in IDP location or sphericity that were likely related to between-manufacturer differences in MRI scanners.

Intra-class correlation coefficients (ICCs) were estimated for pairs of IDP vectors (N=8), each vector comprising measurements of the same IDP in the same subjects at one of 4 possible scanning sites (Chen et al., 2018; Liljequist et al., 2019). The ICC provides a measure of reliability by quantifying the within-subject similarity of each outcome metric or IDP across different sites.

ICCs were estimated by linear mixed effects modeling of variance components, accounting for between-subject and between-site variance, using the lme4 package in R (Bates et al., 2015). We primarily considered ICCs estimated by modelling site as a fixed effect (“consistent ICC”, ICC(3,1)), but also assessed ICCs estimated by modelling site as a random effect (“absolute agreement” ICC or ICC (2,1)). Similar estimates of ICC by both fixed and random effects models will indicate the absence of a systematic bias due to site effects. We estimated ICCs twice: once using all analysable data from 4 sites, including 1 GE site; and once using only Siemens data from 3 sites. ICC values between 0.5 and 0.8 are generally considered to indicate fair to good reliability, and ICCs greater than 0.8 or 0.9 are indicative of good or very good reliability (Koo and Li, 2016).

To benchmark the between-subject and between-site reliability of each IDP measured using the COVID-CNS protocol, we compared these ICCs from the travelling heads study to comparable ICCs estimated in the UKB enhanced cohort. In this design, healthy middle-aged participants were each scanned twice (with mean between-scan interval = 2.25 y; SD = 0.12) at the same one of 4 possible sites, all using the same manufacturer’s system for multi-modal MRI (Siemens SKYRA 3T). We estimated ICCs between the test and retest IDP measurement vectors for N=8 participants, repeatedly, randomly sampled from the total UKB dataset (N = 2,817; 1000 random samples). This allowed us to define a confidence interval for test-retest reliability of each IDP, estimated with N=8, under designed conditions of minimal site and scanner contributions to variance. As noted, the MRI sequences for COVID-CNS were based on similar or identical sequences for T1, T2 FLAIR, dMRI, swMRI and fMRI as previously used in the UKB Enhancement cohort (Table 1). Hence, we could directly compare test-retest and between-site consistency of IDPs measured in the UKB and COVID-CNS cohorts.

## Results

### Sample

Eight participants (7 F; mean age = 23.5 y; SD = 5.8) were successfully scanned at all four sites, with between-site intervals ranging from 1-14 days.

### T1w and T2-FLAIR images

Quality control of T1w and T2 FLAIR images disclosed no deviations in quality of registration (Fig 1a, S1) across sites or with UK Biobank. T1w SNR and CNR measures from Siemens sites were consistent with the UKB population distributions. However, the GE scanner produced images with higher measures of inverse SNR and CNR (equivalent to lower SNR/CNR) than other sites for all subjects (*P*<0.05) (**Figure 1b**). For Siemens sites, across structural IDPs, there was negligible evidence for site-dependent variation in IDP mean values or scaling, and ICC distributions matched those observed in the UK Biobank.

**Figure 1:**
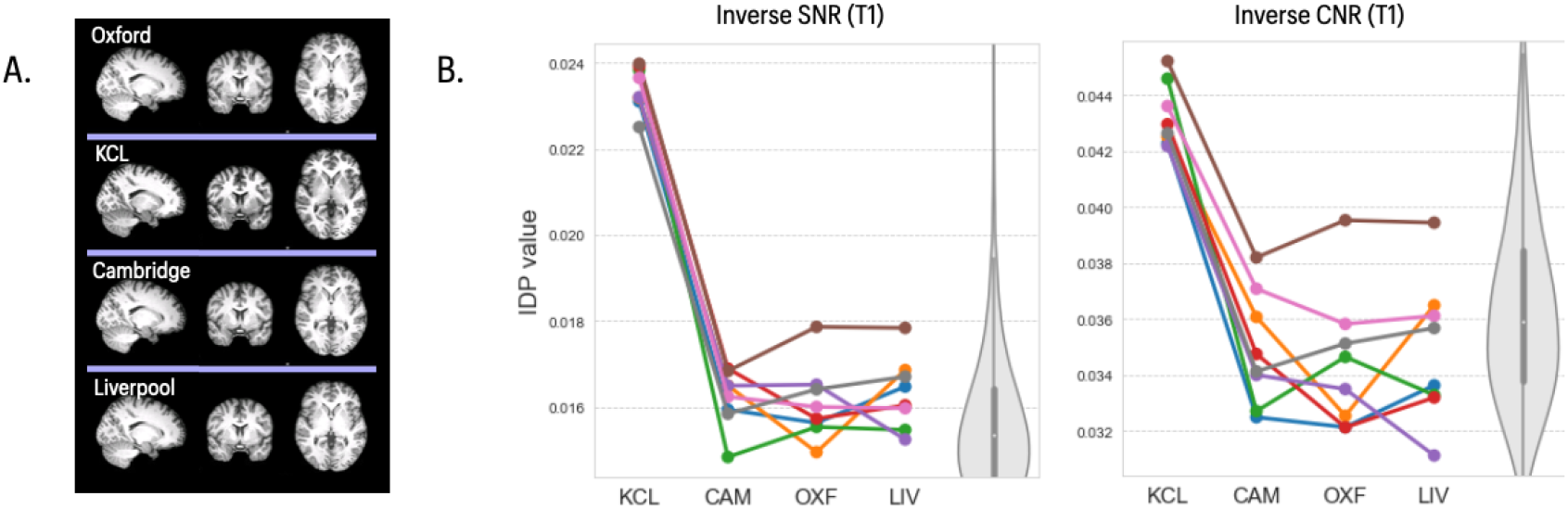
T1 images, inverse SNR and inverse CNR metrics across four sites. **A)** Representative T1 images of the same subject scanned at each of 4 sites in the travelling heads study. **B)** *left panel*, plots of inverse signal-to-noise ratio (iSNR) for 8 subjects (coloured lines) scanned at each of 4 sites (x-axis labels); *right panel*, plots of inverse contrast-to-noise ratio (iCNR) for the same subjects and sites. The grey violin plots in both panels indicate the expected distributions of T1 iSNR and iCNR, respectively, in the UK Biobank reference dataset. The iSNR and iCNR metrics are comparable across Siemens sites (Cambridge, Oxford, Liverpool) and aligned with the UKB benchmark distribution. Both iSNR and iCNR are higher for the GE site (KCL) (P < 0.05), indicating relatively lower SNR and CNR.

Morphometric IDPs, measuring regional volumes and surface areas, showed limited evidence for site-dependent variations in their mean values for Siemens scanners (repeated measures ANOVA; FDR = 5%, **Figure 2a**), and no evidence of significant between-site differences in scaling (Mauchy’s test for sphericity, *P* > 0.05). The GE scanner site had an impact on IDP mean value for a subset of these IDPs. However, consistency across all sites, measured by ICCs, was generally very good for these IDPs (mean ICC >0.9) and did not differ from ICC measures of test-retest consistency in the UKB dataset (**Figure 2b**). Similar results were observed for regional cortical thickness IDPs derived from T1w and T2-FLAIR data. There was some regional variability in between-site (and test-retest) reliability of cortical thickness, but ICCs were typically indicative of good to very good reliability (mean ICC ∼ 0.8), matching those observed in UK Biobank.

**Figure 2:**
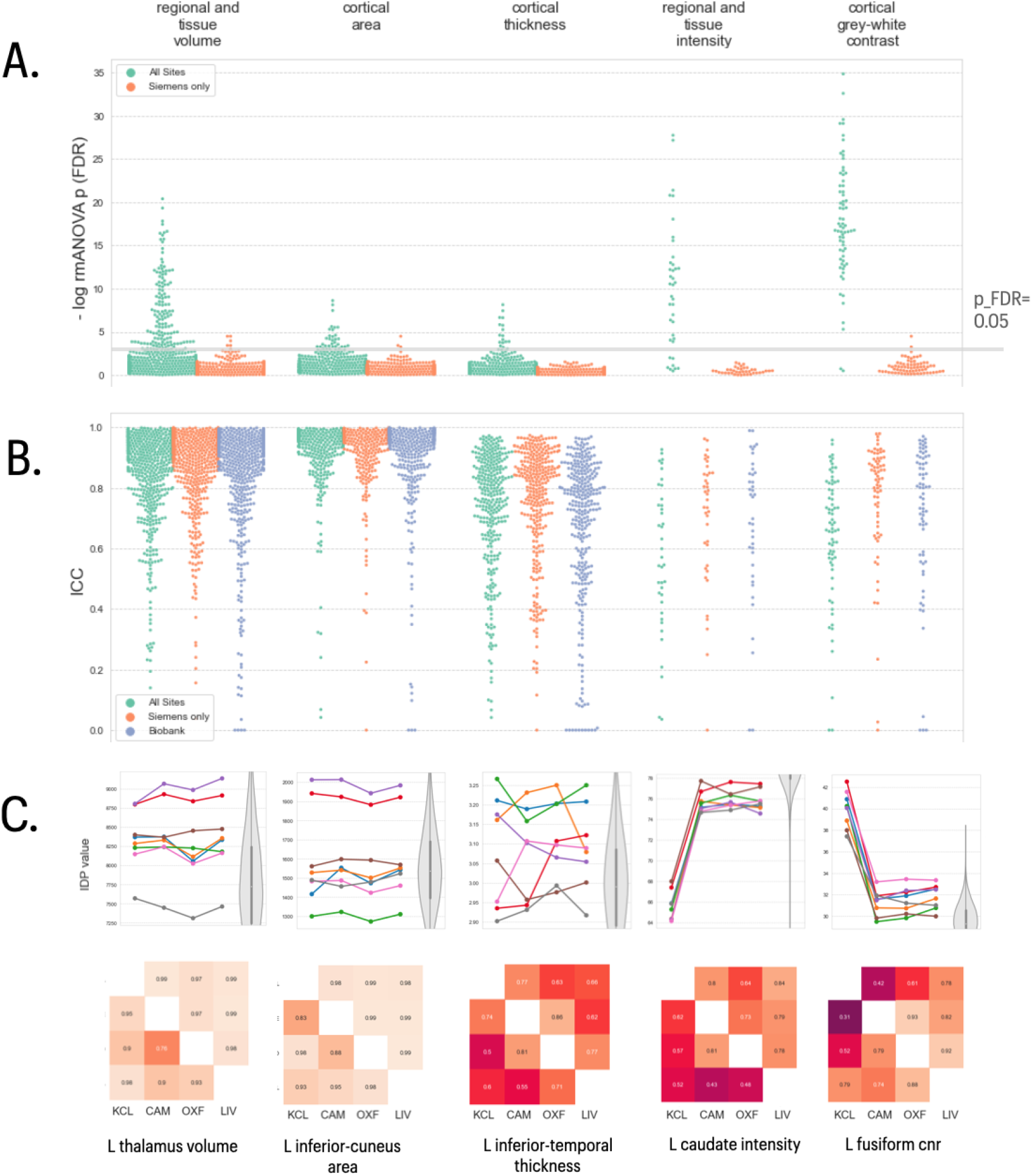
Statistical results for five classes of structural MRI-derived phenotypes. In the top two panels, each column represents results for a different class of IDP, from left to right: regional and tissue volumes, cortical area, cortical thickness, regional and tissue intensity, and cortical grey-white contrast. **A)** Distribution of log-transformed *P*-values from repeated measures ANOVA testing for a site effect on the mean value of individual IDPs in each class; the solid horizontal line represents the P-value equivalent to FDR = 5%. Green dots represent IDPs fitted to the ANOVA model including data from all four sites; orange dots represent P-values for each IDP fitted to the ANOVA including only data from the three Siemens sites (Cambridge, Oxford, Liverpool). There are more significant between-site differences in mean IDPs, across all 5 classes, when the GE data from KCL are included in the analysis **B)** Swarm plots showing distribution of intra-class correlation coefficients (ICCs) for the same IDPs, estimated for each pair of all 4 sites (green points), for each pair of the three Siemens sites (orange points) and for comparable test-retest data drawn from the UKB cohort (blue points). Between-site reliability was generally high for all IDP classes compared to the UKB benchmark, whether or not GE data was included in the analysis. **C)** Each column represents finer-grained results for representative IDPs from each class of IDP: from left to right, left thalamus volume, left precuneus area, left inferior temporal cortical thickness, left caudate intensity and left fusiform CNR. *Top row*, plots of each IDP for 8 subjects (coloured lines) scanned at each of 4 sites (x-axis labels); the grey violin plots indicate the distributions of the corresponding IDP in the UK Biobank reference dataset. *Bottom row*, correlations between each pair of sites for each IDP: upper triangle, Pearson’s correlations; lower triangle, Spearman’s correlations.

Tissue intensity and grey-white contrast IDPs were again consistent across Siemens sites, but often showed significant differences at the GE site. There were significant differences in mean tissue intensity and grey-white contrast for 79% and 97%, respectively, of regional IDPs (RM ANOVA, FDR=5%). Grey-white contrast measured on the GE data was generally lower than in the Siemens data, reflecting the effects observed in the global SNR and CNR measures (Figure 1b). Between-site reliability for these IDPs across the 3 sites using Siemens scanners was slightly higher (mean ICC = 0.69, SD = 0.24) than between-site reliability across all 4 sites (mean ICC = 0.61 SD = 0.23), compared to a UK Biobank mean ICC of 0.66 (SD = 0.17).

White matter hyper-intensity volumes (WMHs) derived from T2-FLAIR images of the healthy young adults scanned in the travelling heads study were typically low, as expected in this age range (21-37 y). However, there were significant mean differences between sites in both deep and periventricular WMH volumes (RM ANOVA; FDR = 5%), due to greater WMH volumes in the GE data, with correspondingly lower levels of between-site reliability (**Figure 3**). There were no significant mean differences between Siemens sites in deep or periventricular WMH volumes and between-site reliability for the 3 Siemens sites was very good (ICC = 0.95, sd=0.01), comparable to test-rest reliability in the UKB data (ICC = 0.90, sd=0.06), and greater than between-site reliability over all 4 sites in the travelling heads data (ICC = 0.51, sd=0.12). These findings are somewhat unsurprising given that the software tool for WMH measurement (BIANCA) was trained on data collected from the Siemens MRI protocol. When adequate training data are available from the GE protocol, and in older subjects where higher WMH volumes are expected, it will be important to retrain the BIANCA algorithm on both Siemens and GE data and this may improve consistency of WMH IDPs across scanners from the different manufacturers (Bordin et al., 2020).

**Figure 3:**
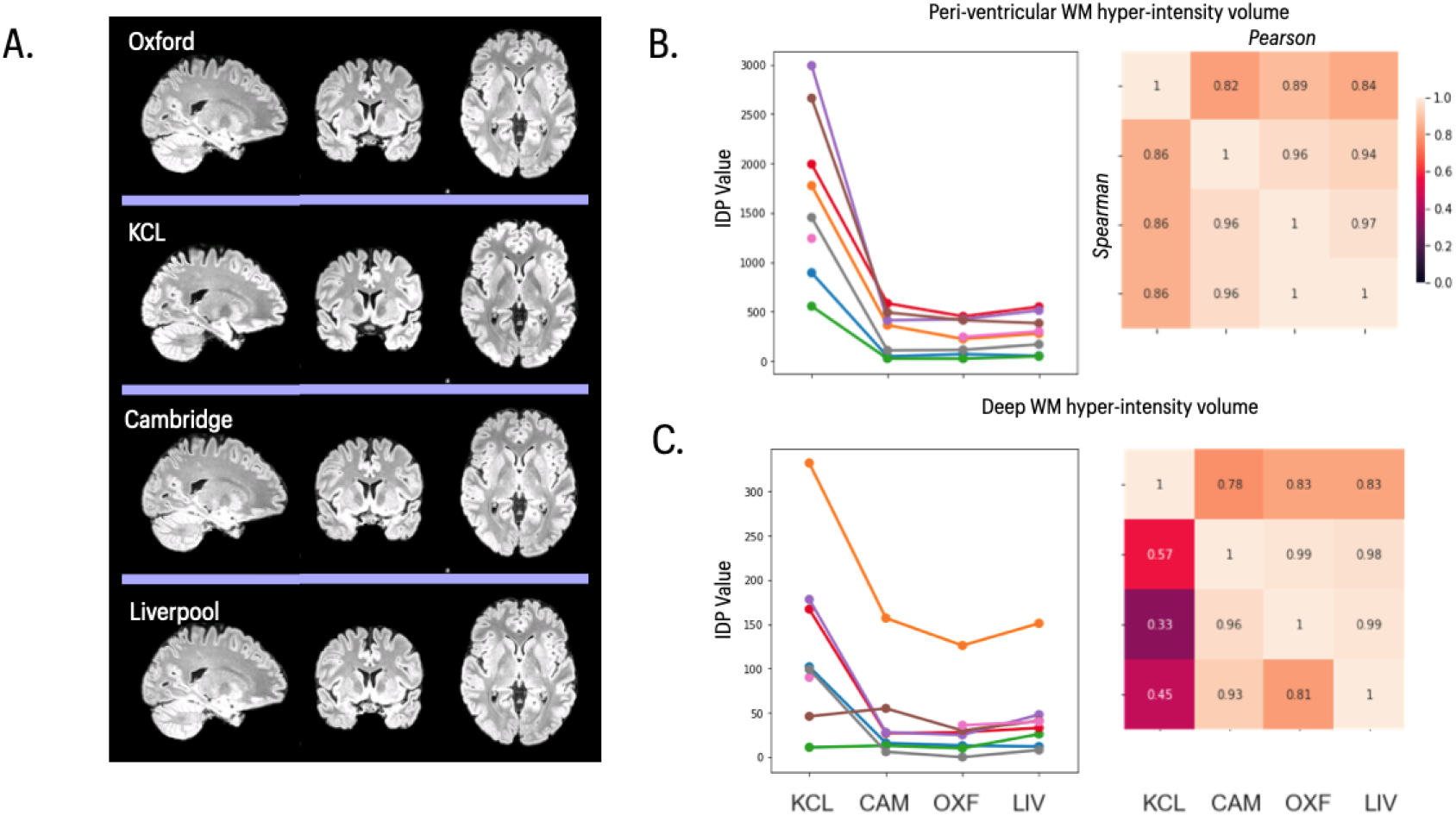
T2 FLAIR images and statistical results for T2-derived IDPs. **A)** Representative T2 FLAIR images of the same subject scanned at each of 4 sites in the travelling heads study. **B)** *left panel*, peri-ventricular white matter hyperintensity volume for 8 subjects (coloured lines) scanned at each of 4 sites (x-axis labels); *right panel*, correlations between each pair of sites. **C)** *left panel*, deep white matter hyperintensity volume for 8 subjects (coloured lines) scanned at each of 4 sites (x-axis labels); *right panel*, correlations between each pair of sites. In both **B)** and **C)**, the upper triangle of the matrix shows Pearson’s correlations and the lower triangle shows Spearman’s correlations; and both IDPs were estimated using BIANCA.

### Susceptibility Weighted Imaging

We assessed regional estimates of T2^*^ signal decay and quantitative estimates of susceptibility (QSM) derived from the swMRI images. There was limited evidence of site-specific variation in IDP means or scaling (Fig 4a). Estimates of regional T2^*^-had poor between-site reliability across all 4 sites in the travelling heads data (mean ICC = 0.34, sd=0.24) (**Figure 4b**). QSM-derived IDPs had generally better between-site reliability (All sites: ICC = 0.67, sd=0.13; Siemens only: ICC = 0.76, sd=0.14), comparable to good-very good test-retest reliability in the UKB data (ICC=0.66). Lower reliability was observed for QSM IDPs measured in smaller subcortical structures (amygdala, nucleus accumbens) in both travelling heads and UKB datasets.

**Figure 4:**
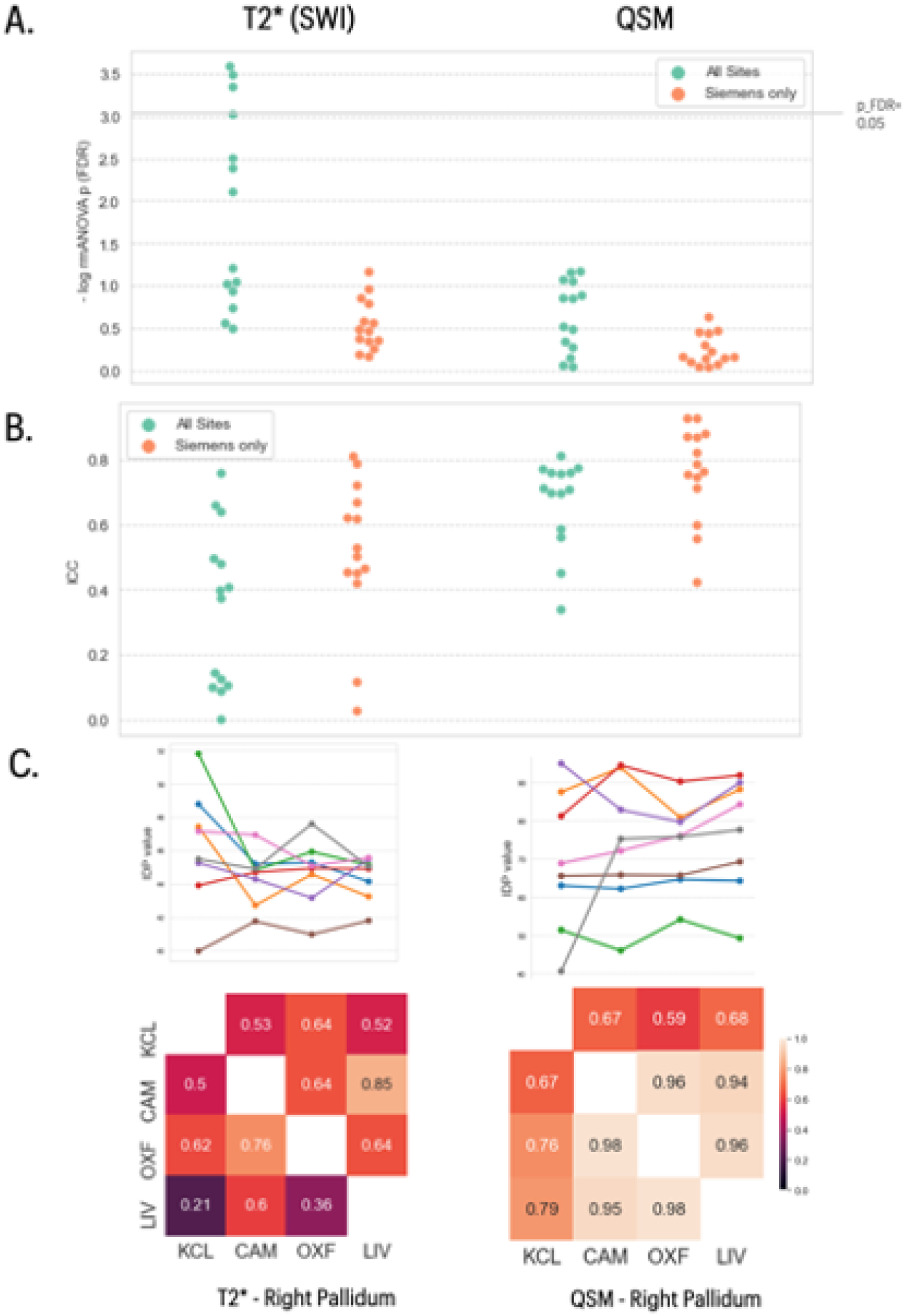
Statistical results for SWI-derived IDPs. In the top two panels, the left column shows data for 14 IDPs derived from T2^*^ data and the right column shows data for 14 IDPs derived from QSM data. **A)** Distribution of log-transformed *P*-values from repeated measures ANOVA testing for a site effect on the mean value of individual IDPs in each class; the solid horizontal line represents the P-value equivalent to FDR = 5%. Green dots represent IDPs fitted to the ANOVA model including data from all four sites; orange dots represent P-values for each IDP fitted to the ANOVA including only data from the three Siemens sites (Cambridge, Oxford, Liverpool). There are more significant between-site differences in mean IDPs when the GE data from KCL are included in the analysis **B)** Swarm plots showing distribution of intra-class correlation coefficients (ICCs) for the same IDPs, estimated for each pair of all 4 sites (green points), and for each pair of the three Siemens sites (orange points). **C)** Each column represents finer-grained results for representative IDPs from each class of IDP: from left to right, T2^*^ right pallidum, QSM right pallidum. *Top row*, plots of each IDP for 8 subjects (coloured lines) scanned at each of 4 sites (x-axis labels). *Bottom row*, correlations between each pair of sites for each IDP: upper triangle, Pearson’s correlations; lower triangle, Spearman’s correlations.

### dMRI

Diffusion weighted images were successfully acquired and analysed at all sites. Visualisation and basic QC metrics showed consistent image quality across sites. IDPs corresponding to multiple diffusion parameters (FA, MO, MD, ICVF, ISOVF and OD), were estimated regionally for each of multiple white matter tracts. As for other modalities some IDPs showed evidence for site-specific differences in means, driven by the GE site (**Fig 5a**). Overall, there was good to very good between-site reliability (mean ICCs > 0.7), matching those observed in the UKB (**Fig 5b**). The GE site showed limited consistency with other sites for WM tract FA, diffusivity and ISOVF, reducing ICCs for these categories of IDPs.

**Figure 5:**
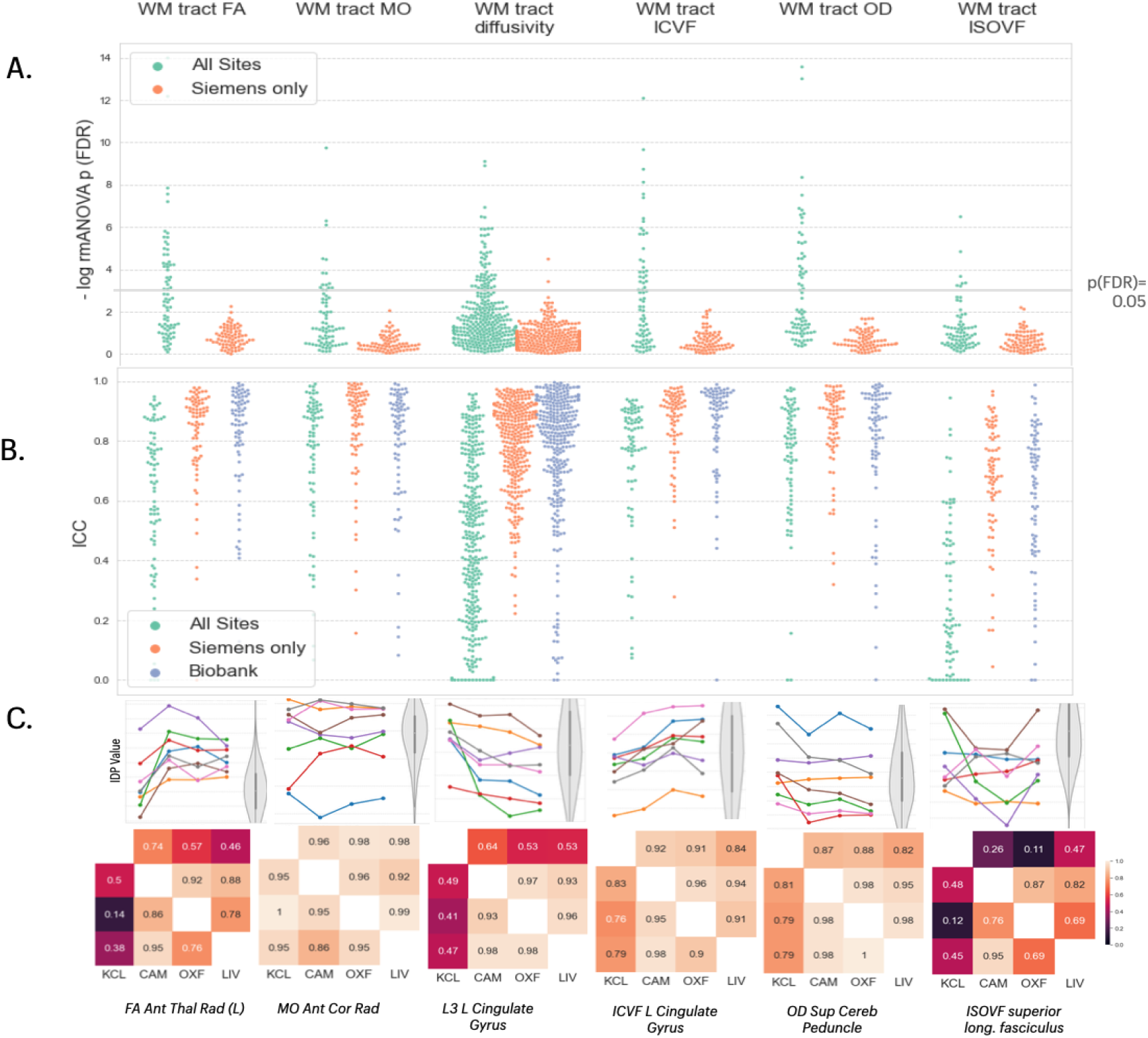
Statistical results for five classes of dMRI-derived phenotypes. In the top two panels, each column represents results for a different class of IDP, from left to right: Fractional white matter (WM) tract FA, WM tract MO, WM tract diffusivity, WM tract ICVF, WM tract OD and WM tract ISOVF. **A)** Distribution of log-transformed *P*-values from repeated measures ANOVA testing for a site effect on the mean value of individual IDPs in each class; the solid horizontal line represents the *P*-value equivalent to FDR = 5%. Green dots represent IDPs fitted to the ANOVA model including data from all four sites; orange dots represent *P*-values for each IDP fitted to the ANOVA including only data from the three Siemens sites (Cambridge, Oxford, Liverpool). There are more significant between-site differences in mean IDPs, across all 5 classes, when the GE data from KCL are included in the analysis **B)** Swarm plots showing distribution of intra-class correlation coefficients (ICCs) for the same IDPs, estimated for each pair of all 4 sites (green points), for each pair of the three Siemens sites (orange points) and for comparable test-retest data drawn from the UKB cohort (blue points). Between-site reliability was generally high for all IDP classes compared to the UKB benchmark when only Siemens sites were included in the analysis. **C)** Each column represents finer-grained results for representative IDPs from each class of IDP: from left to right, FA right anterior thalamic radiation. MO left corona radiata, L3 left cingulate gyrus, ICVF left cingulate gyrus, OD superior cerebellar peduncle, and ISOVF superior longitudinal fasciculus. *Top row*, plots of each IDP for 8 subjects (coloured lines) scanned at each of 4 sites (x-axis labels); the grey violin plots indicate the distributions of the corresponding IDP in the UK Biobank reference dataset. *Bottom row*, correlations between each pair of sites for each IDP: upper triangle, Pearson’s correlations; lower triangle, Spearman’s correlations.

### fMRI

Resting fMRI was successfully acquired at all sites . There were no significant between-site differences in mean tSNR (before or after ICA-based processing with FIX), indicating similar levels of signal quality across all sites, with QC metrics commensurate with those observed in the UKB data (**Figure 6**). As individual functional connectivity (FC) IDPs reflecting pairwise connectivity did not show a high level of reliability across sites, we assess 6 modes of variation of FC network connectivity shown to be reliable in UKB (Elliott et al., 2018). We also assess individual node amplitudes. These IDPs in general did not show site-specific variations in mean or scaling (Fig 6A). Between-site reliability was low for node amplitudes (All sites: mean=0.36 sd=0.17; Siemens mean=0.55 sd=0.19), but comparable to the UKB (mean=0.48 sd=0.27). The 6 RSN connectivity modes showed very good reliability, with mean ICC = 0.67 (sd=0.18) for all sites and ICC=0.75 (sd=0.25) for Siemens sites, compared to the excellent reliability seen in the UBK (mean ICC=0.89, sd=0.11).

**Figure 6.**
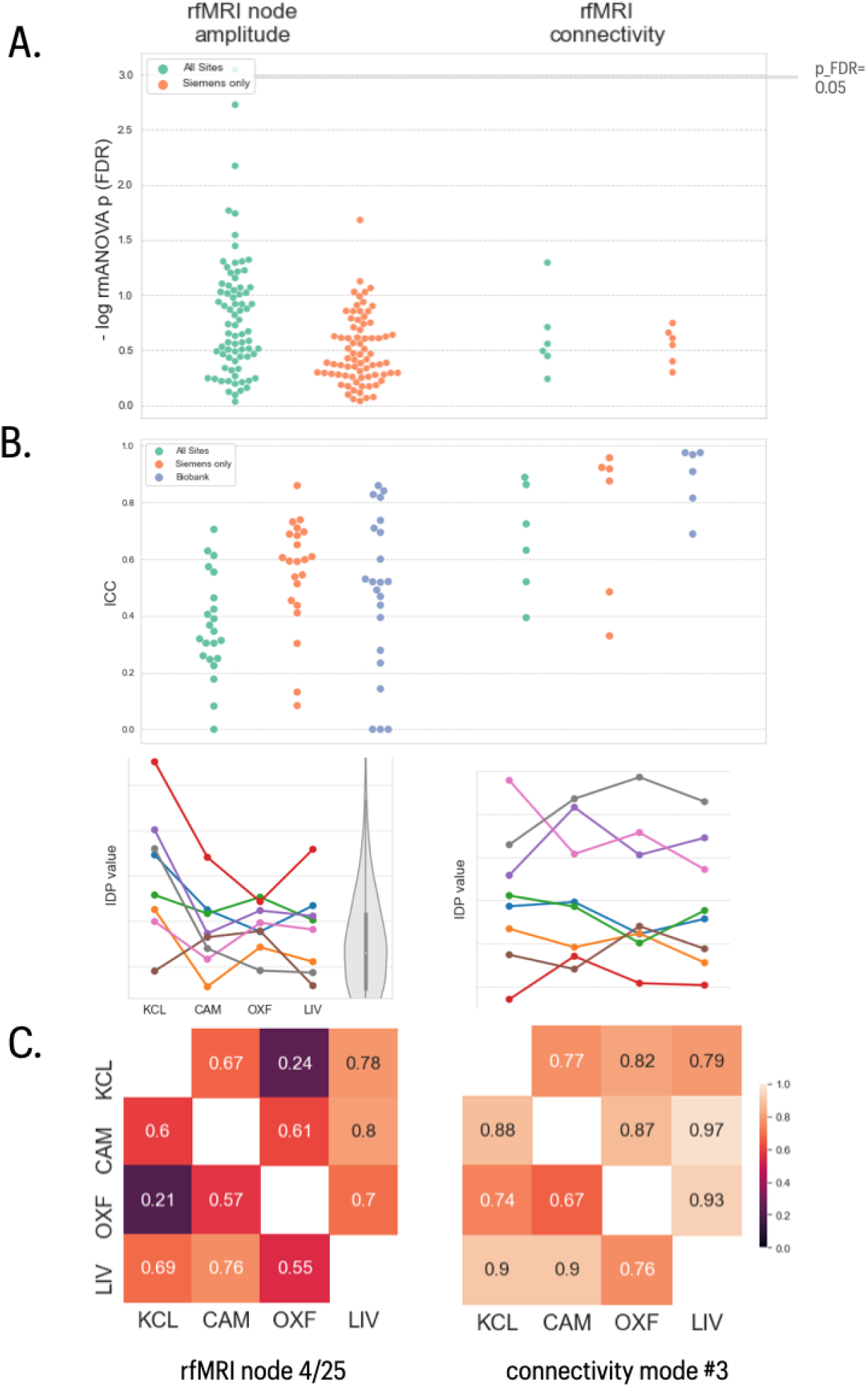
fMRI data quality and IDP summaries. The two columns show data on fMRI node amplitude and fMRI connectivity IDPs. Both represent IDPs derived from 25- and 100-node ICA-based parcellations. The fMRI connectivity IDPs represent 6 modes of variation across the functional connectivity network matrices derived from both parcellations. **A**) Distribution of log-transformed *P*-values from repeated measures ANOVA testing for a site effect on the mean value of individual IDPs in each class; the solid horizontal line represents the P-value equivalent to FDR = 5%. Green dots represent IDPs fitted to the ANOVA model including data from all four sites; orange dots represent P-values for each IDP fitted to the ANOVA including only data from the three Siemens sites (Cambridge, Oxford, Liverpool). **B)** Swarm plots showing distribution of intra-class correlation coefficients (ICCs) for the same IDPs, estimated for each pair of all 4 sites (green points), for each pair of the three Siemens sites (orange points) and for comparable test-retest data drawn from the UKB cohort (blue points). Between-site reliability was generally high for all IDP classes compared to the UKB benchmark, whether or not GE data was included in the analysis. **C)** Each column represents finer-grained results for representative IDPs from each class of IDP: from left to right, fMRI node 4/25 and connectivity mode #3. *Top row*, plots of each IDP for 8 subjects (coloured lines) scanned at each of 4 sites (x-axis labels); the grey violin plot indicates the distribution of the corresponding IDP in the UK Biobank reference dataset. *Bottom row*, correlations between each pair of sites for each IDP: upper triangle, Pearson’s correlations; lower triangle, Spearman’s correlations.

### Arterial spin labelling

For both the single PLD sequence (acquired on all sites) and the multi-PLD sequence (acquired on the three Siemens sites only), we assessed estimates of grey and white matter mean perfusion. Due to acquisition challenges, ASL was not successfully acquired at all sites. There was no evidence of between-site mean differences in estimated perfusion. Between-site reliability for the single PLD sequence was poor (ICC = 0.35; Siemens sites only ICC=0.22). The Siemens-only multi PLD sequence had fair reliability (ICC = 0.53) (**Figure 7**).

**Figure 7.**
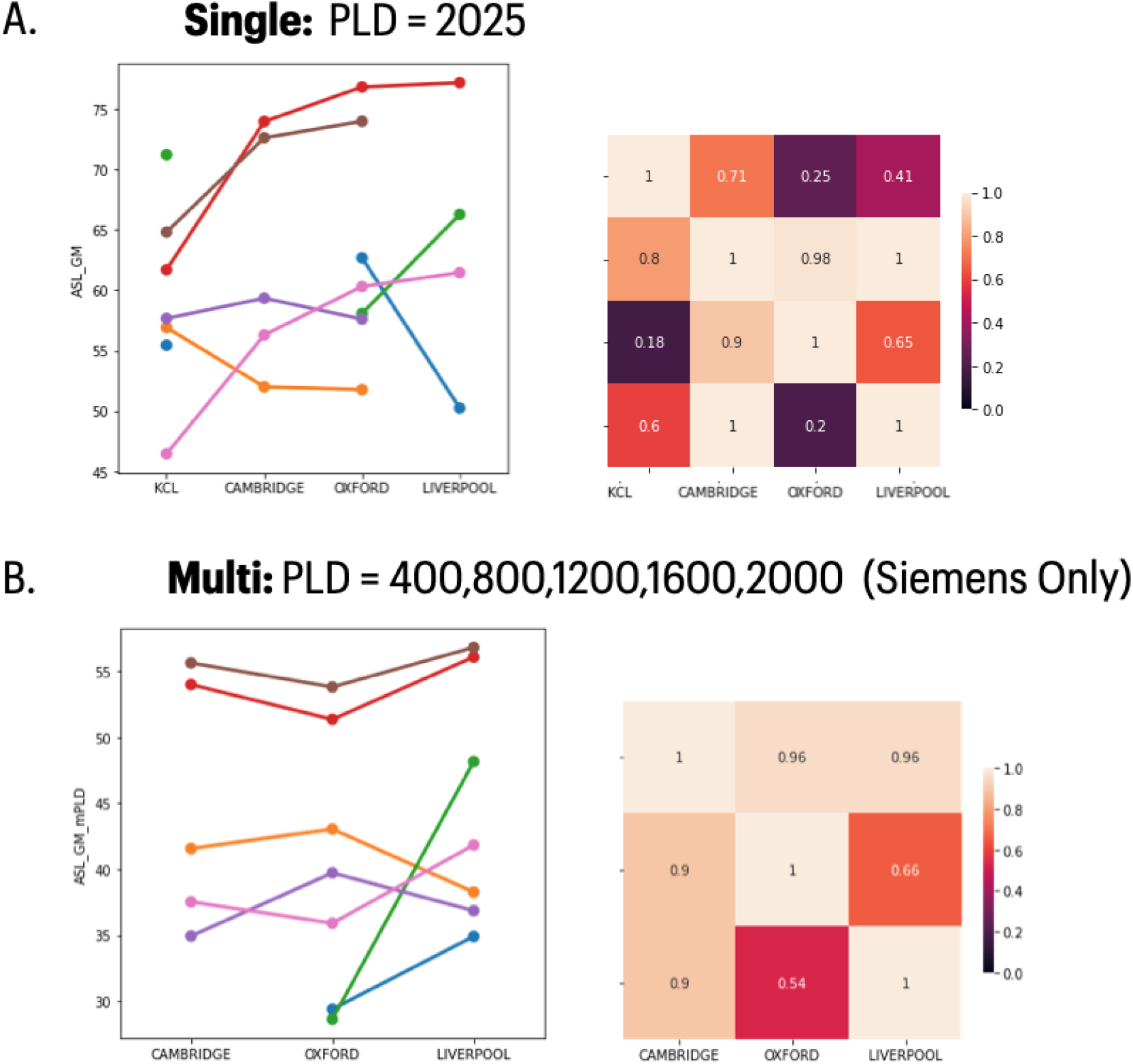
ASL data IDP summaries. **A)** Grey matter mean perfusion data for the single post-label delay (PLD) sequence used across all sites. **B)** Grey matter mean perfusion data for the multi-PLD sequence available only on the Siemens sites. Raw data is plotted to the left; the cross-site correlation matrices to the left (upper triangle, Pearson’s correlation; lower triangle, Spearman’s correlation).

## Discussion

This study provides a detailed investigation of the reliability of multi-modal IDPs for the multi-site COVID-CNS project. This work provides one of the broadest surveys of the reliability of multi-modal neuroimaging measures to date. For the COVID-CNS project, it provides insights that can guide the design of harmonisation strategies for the project. More broadly, the study is of relevance to the expanding number of studies utilising multi-modal imaging protocols derived from the UK Biobank, including a number of additional studies focused on the neurological impact of COVID-19 (Douaud et al., 2021; Raman et al., 2021). Overall, our results demonstrate generally good to excellent levels of between-site reliability of imaging derived phenotypes estimated across a wide range of brain MRI modalities in data collected from 4 UK sites participating in a national COVID research consortium. In particular, the 3 sites using Siemens PRISMA platforms reliably estimated from repeated measures on participants sampled from the UKB database. When the site from a different scanner manufacturer (GE) was included, certain IDP classes were less reliable. These results give confidence that large, multi-site COVID imaging studies can be used to expand the cohort sizes of COVID neuroimaging studies.

Variability in IDPs across sites may be induced by variation in the contrast obtained by specific sequences and scanner setups or technical variation in signal levels, scaling, or SNR. Travelling heads studies provide a powerful means by which to detect site-specific variations in these features in advance of multi-site population studies. In a healthy-participant travelling heads study, ICC depends on intrinsic inter-subject variation in the travelling heads cohort to drive measures of reliability. As such, ICC may be an imperfect measure to compare IDPs, as between-subject variability may not reflect the observed effect size in the condition of interest for individual IDPs (e.g. neurological effects of COVID). Nevertheless, ICCs are valuable when it is expected that clinical effect sizes will be on the approximate scale of individual variation, and for comparison to other datasets (e.g. UKB). While N=8 provides limited statistical power for the identification of subtle differences across sites in individual IDPs, here it was able to provide an overall pattern of results indicating that there will not be substantial loss of statistical power when introducing new sites.

### Reliability of multi-modal, multi-site MRI measurements

The between-site reliability for the 3 sites using Siemens PRISMA platforms allow us to evaluate which MRI sequences and IDPs were most (and least) operationally and statistically reliable under the best-case scenario of nearly identical scanners at multiple sites. The most reliably collected MRI sequences were T1w, T2-FLAIR and dMRI; the least reliably collected MRI sequence was ASL (N=6). This is perhaps unsurprising given the relative novelty of these ASL sequences, which are well-established for research at specialised centres but had not previously been used for large-scale clinical studies at all sites participating in the travelling heads study.

The most reliably estimated IDPs were geometric grey matter phenotypes (cortical volume, surface area, thickness), and white matter microstructural phenotypes (FA, ODI etc). Between-site reliability for these two classes of IDPs was excellent in the travelling heads data and comparable to the ceiling level of test-retest reliability of the same classes of IDP in the UKB dataset. Less reliably estimated IDPs were typically derived from the less reliably collected ASL data; but all other classes of IDP had good-to-excellent levels of both between-site and test-rest reliability. It was notable that the ICCs for between-site and test-retest reliability were positively correlated across all IDPs derived from Siemens data in the travelling heads and UKB studies, indicating that some IDPs are inherently more robust to both between-site and within-subject sources of variation. This may have implications for the power to detect case-control differences in clinical studies using this set of multi-modal MRI sequences. For example, if there were comparable effect sizes and sample sizes, T1w, T2-FLAIR and dMRI-derived IDPs will clearly have greater power to detect case-control differences by virtue of their lower (between-site) variability.

### Between-manufacturer reliability of multi-modal MRI measurements

The GE platform increased between-site variability for many classes of IDP, showing significant differences in mean and reductions in ICC. Clearly, this increased between-site variability was driven by differences in MRI sequences and data between Siemens and GE scanner platforms. Despite careful preparatory alignment of the GE sequences to approximate as closely as possible the parameters of the Siemens sequences, there were some irreducible differences between Siemens and GE protocols due to the hardware constraints of differently manufactured scanners. Tissue contrast metrics, like grey/white matter contrast, were particularly sensitive to the difference between Siemens and GE sites, whereas geometric grey matter IDPs were generally more robust. The reliability of white matter hyperintensity volume estimation was notably poor when GE data were included in the analysis, but this may be at least partly attributable to the fact that WMH volumes were estimated in healthy young adults (not usually expected to have any WMHs) using a software tool that had been trained on Siemens-only data. Further training of WMH segmentation tools on data acquired from GE as well as Siemens platforms in older subjects would likely improve the reliability of this key marker of inflammation-related changes in white matter.

For a nationally-scaled study of post-COVID patients, these data clearly point to a trade-off between increasing recruitment rates (and ultimately sample size) by including sites using scanners supplied by different manufacturers *versus* maximising between-site reliability (and thus reducing spurious sources of variability) by restricting sites to those that are using scanners supplied by the same manufacturer. Geographical differences in the incidence of COVID, and in operational capacity for research studies under pandemic conditions, motivated formation of a large and nationally representative network of scanning sites. We considered that the generally good-to-very good levels of reliability for most IDPs across all sites in this pilot study were sufficient to support this more inclusive strategy of using sites with either Siemens or GE scanners, with the caveat that this will entail loss of power to detect case-control differences in terms of IDPs derived from ASL and other modalities which were most difficult to harmonize between manufacturers. Between-site offsets in the mean and scaling of IDP values could be corrected statistically post hoc by standard harmonisation or modelling methods such as COMBAT(Da-ano et al., 2020) or Generalised Additive Modelling (Dinga et al., 2021), so long as certain sampling requirements for patients and controls can be achieved at individual sites.

### Methodological issues

It is a strength of this study that we have assessed reliability across a wide range of MRI modalities and imaging-derived phenotypes, using data collected from different MRI systems and at different sites. It is also a strength that we have been able to benchmark between-site reliability for the majority of IDPs against comparable estimates of test-retest reliability in the UKB data.

However, sample size for the travelling heads study was small, meaning that results were potentially vulnerable to the effects of 1 or 2 outlying observations and confidence intervals were generally wide. We made best efforts, under the pragmatic constraints of urgently responding to a public health crisis, to align GE and Siemens sequences prior to data acquisition. However, we cannot claim that the between-manufacturer reliability results are optimised or would be unimprovable by future, more intensive work on Siemens-like sequences for sites using scanners supplied by GE or other manufacturers to align with UKB and C-MORE standards for COVID neuroimaging. The results also indicate strong prospects for the wider integration of COVID-related clinical neuroimaging data, particularly when sequences are reasonably aligned across studies.

### Conclusion

These results represent a realistic guide to the generally acceptable to excellent levels of between-site reliability that are immediately attainable for multi-modal MRI across a national network of collaborating sites using different scanner platforms. The UK Biobank multimodal imaging protocols, which we have translated here to other sites and scanner models, presents an attractive suite of protocols for new studies to consider using to ensure strong reusability of data.

## Data Availability

All data produced in the present study will be made openly available when the paper has been peer-reviewed.

## Data sharing statement

Individual de-identified participant data will be shared through a secure online platform in support of peer-reviewed publication of this report. The main UK Biobank brain MRI analysis pipeline is available at https://www.fmrib.ox.ac.uk/ukbiobank/. Modified or additional scripts and support data for the analyses performed in this study will be made available from covidcns.org.

## Acknowledgements

We thank the volunteers who participated in the travelling heads study and the radiography staff who collected data at all four sites. We thank Fraunhofer MEVIS, Bremen, Germany, for provision of the 3D-GRASE ASL sequence

